# COVID-19 Vaccination is Associated with Decreasing Cases, Hospitalizations, and Deaths Across Age Groups and Variants over 9 months in Switzerland

**DOI:** 10.1101/2022.02.03.22270396

**Authors:** Jean-Luc Jucker

## Abstract

Recent studies assessing COVID-19 vaccine efficacy at the population level found counterintuitive results, such as *positive* associations between vaccination and infections or deaths. These ecological studies have limitations, including too short observation periods, focusing on infections, and not controlling for age groups and dominant variants. The current study addresses these limitations by investigating the relations between vaccination and COVID-19 cases, hospitalizations, and deaths over a longer period (9½ months) while also considering age groups (from 10 to 80+ years old) and variants (Alpha and Delta), utilizing data from Switzerland. Results suggest that vaccination is negatively related to cases overall and in all cantons of Switzerland, and that vaccination is negatively related to hospitalizations and deaths from 50 years old. Furthermore, vaccination is a significant predictor of cases, hospitalizations, and deaths while holding the effects of age and dominant variant constant.

## Introduction

Vaccination is as of yet the main strategy to control the SARS-CoV-2 (COVID-19) pandemic and it is therefore important to regularly assess its efficacy. Several recent ecological studies have attempted to do so, and reported counterintuitive results [1-3]. For example, Fukutani et al. [2] found that in 60 and 37 countries, vaccination is *positively* correlated with COVID-19 infections and deaths, respectively. Likewise, Subramanian and Kumar [3] reported that increases in COVID-19 cases are unrelated to vaccination across 68 countries as well as in most United States counties^1^.

Ecological approaches have limitations, including that they cannot provide evidence for causality at the individual level [5-6]. Yet the positive associations between vaccinations and infections found in the above-mentioned studies are hard to explain. Björk et al. [5] showed that differences in population size should be taken into account, and Melton and Sinclair (preprint here) found different effects when adding median age (though not age groups) to their analyses. These studies, however, also have limitations. None of them analyzes data by age groups (arguably an essential determinant), most focus on infections or deaths (excluding hospitalizations), and most also use short observation periods (from 7 days to 3½ months, see also Backhaus, preprint [here](https://osf.io/preprints/socarxiv/dzk4j/)). Furthermore, none compared vaccine efficacy across COVID-19 variants.

The aim of the current ecological study is to address these limitations by investigating the relations between vaccination and COVID-19 cases, hospitalizations, and deaths over a significantly longer period (9½ months), while also taking into account age groups and variants. For this, it utilizes data from Switzerland.

### Data and Methods

Data from the Swiss Federal Office of Public Health (FOPH), which are available here, were downloaded and processed on 10 January 2022. Several files were merged in Python to obtain a single dataset with the following main variables: (1) the incidence of laboratory confirmed COVID-19 cases (hereafter, Cases); (2) the incidence of COVID-19 related hospitalizations (Hospitalizations); (3) the incidence of COVID-19 related deaths (Deaths); (4) the percentage of fully vaccinated persons in the population^2^ (Vaccination); (5) the current dominant COVID-19 variant (Variant), which was either B.1.1.7 (hereafter, Alpha) or B.1.617.2 (Delta).

The incidence for Cases, Hospitalizations, and Deaths, was computed per 100,000 inhabitants per week. The dominant Variant was defined as the variant representing the highest percentage of all COVID-19 related infections at any given time (rolling 7-day average).

These data covered the period from 17 January 2021 to 31 October 2021 (9½ months or 42 weeks), with one measurement per week for each variable^3^. The reason to choose this period was that it represents a wide range of vaccinated persons (from 0% fully vaccinated persons on 17 January 2021 to 64.02 fully vaccinated persons on 31 October 2021)^4^ and two subsequent dominant Variants (Alpha for the first 23 weeks, Delta for the last 19 weeks^5^). Furthermore, except for Variant, these data were available at several levels, including region (Switzerland^6^ overall, and the 26 cantons separately) and Age Group (10-19, 20-29, 30-39, 40-49, 50-59, 60-69, 70-79, and 80+ years old^7^, hereafter Age Group), so that the rates of Vaccination, Cases, Hospitalizations, and Deaths could be used in analyses accordingly.

To analyze these data, we first computed correlation coefficients between Vaccination and Cases, Hospitalizations, and Deaths for the whole of Switzerland, then separately by region and Age Group, and finally by both Age Group and Variant. We then used multiple linear regression to assess whether Vaccination is a significant predictor of Cases, Hospitalizations, and Deaths while controlling for the effect of Age Group and dominant Variant.

## Results

At the Switzerland level, Vaccination was found to be significantly negatively correlated to Cases (r = -0.396, p < 0.001^8^), not significantly positively correlated to Hospitalizations (r = 0.013, p = 0.669), and borderline significantly negatively correlated to Deaths (r = -0.054, p = 0.086).

When the same analysis was ran by region, more nuanced results were obtained. As one can see in Figure 1a, across all 26 cantons, Vaccination is negatively correlated to Cases, however with coefficients that differ between some cantons (r range = -0.039 to -0.63). In turn, for most cantons, Vaccination is positively correlated to Hospitalizations, although in 7 cantons, including 2 with the more inhabitants (Zürich and Vaud), it is negatively correlated to Hospitalizations, which is in line with [5]. Finally, Vaccination is negatively correlated to Deaths in 16 of the 26 cantons, with the highest positive correlation observed (r = 0.15) in the canton with the least inhabitants (Appenzell Innerrhoden).

**Fig. 1.**
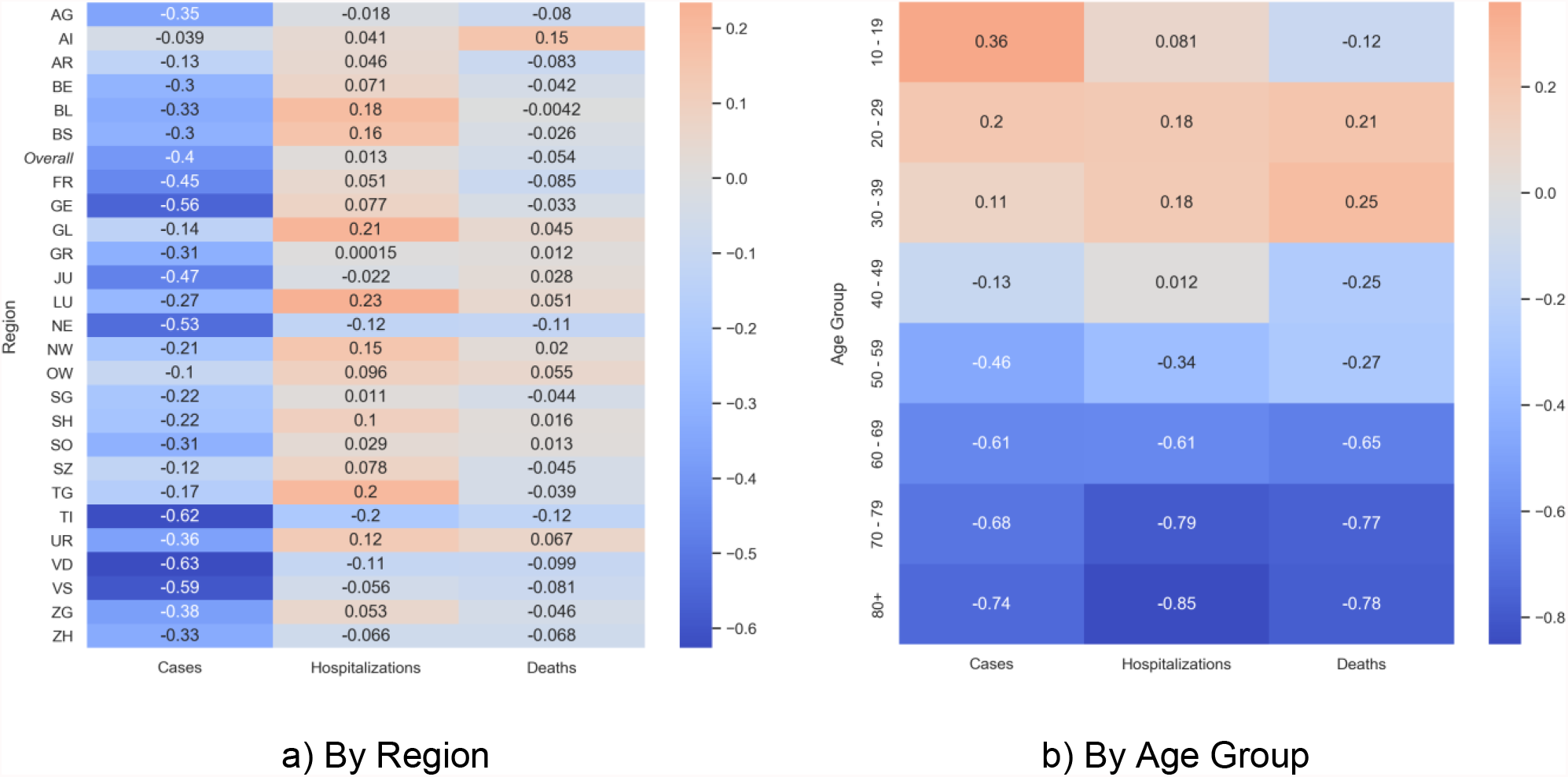
Correlations Between Vaccination and Cases, Hospitalizations and Deaths

The same analysis ran by Age Group revealed another, yet unsurprising trend. Vaccination tends to be positively correlated to Cases, Hospitalizations, and Deaths until approximately 40 years old, and it then becomes more and more negatively correlated to these same variables. Importantly, one can see in Figure 1b that from 60 years old, Vaccination is strongly negatively correlated not only to Cases (r = -0.74), but even more to Deaths (r = -0.78) and Hospitalizations (r = -0.85). The fact that the highest negative correlation observed concern Hospitalizations, which was insignificant at the Switzerland overall, illustrates well the importance of considering age groups separately.

We finally ran correlations grouped by both Age Group and dominant Variant. As one can see in Figure 2, results seem to differ between Variants. For example, across Age Groups, Vaccination is negatively correlated with Alpha Cases, whereas it is positively correlated with Delta Cases. Similar results were obtained for Hospitalizations and Deaths, as one can see in the Supplementary Information.

**Fig. 2.**
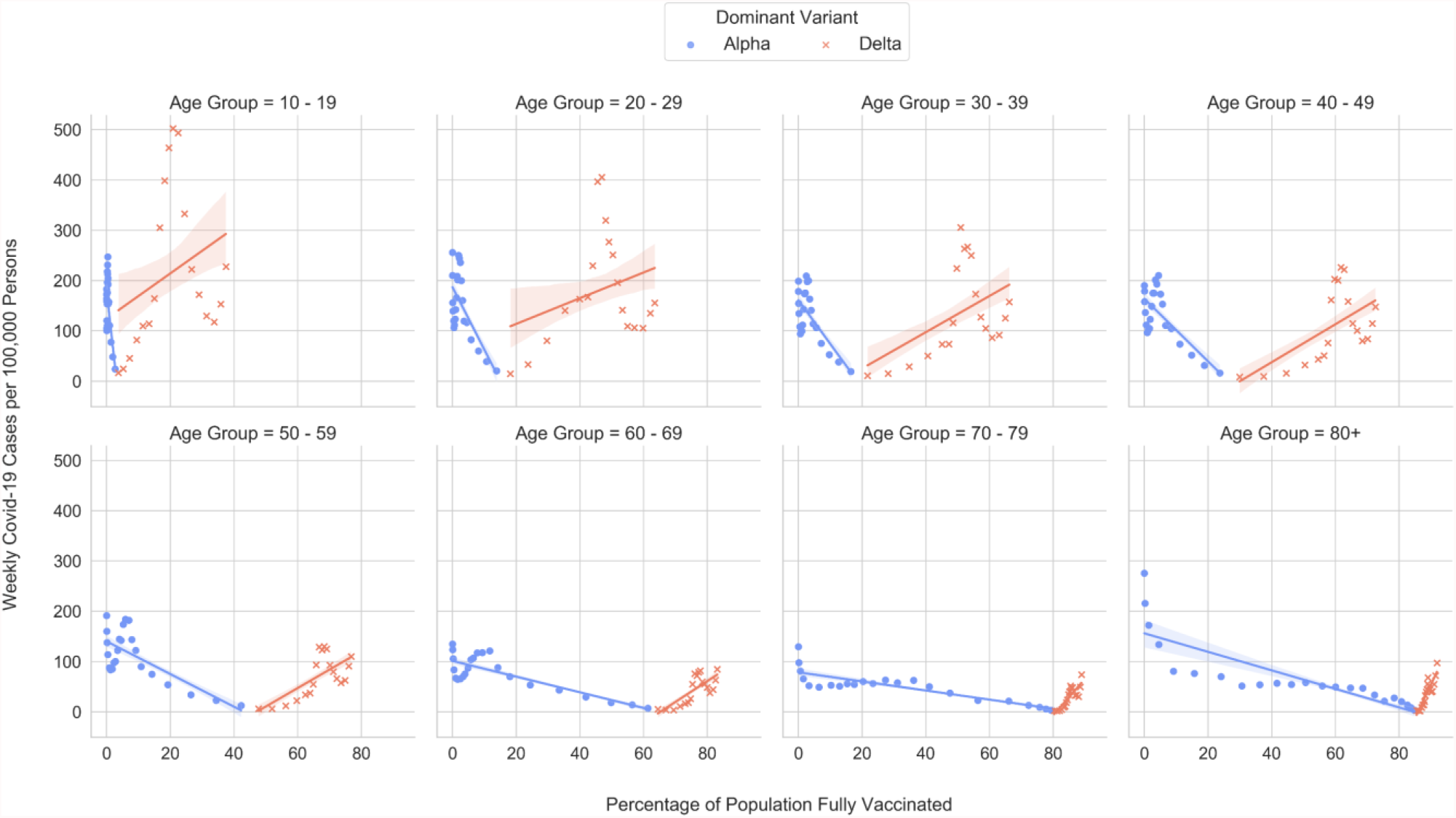
Relationships between Vaccination and Cases by Age Group and Dominant Variant

To better assess the role of vaccination, multiple linear regression was used to predict Cases, Hospitalizations, and Deaths, while controlling for the effects of Age Group and Variant at the Switzerland level. Results are presented in Table 1. As one can see, Variants and most Age Groups are significant predictors of Cases, Hospitalizations, and Deaths. Importantly, Vaccination is also a significant predictor and is associated with a decrease in Cases, Hospitalizations, and Deaths when holding the effect of Age Group and Variant constant.

**Table 1.**
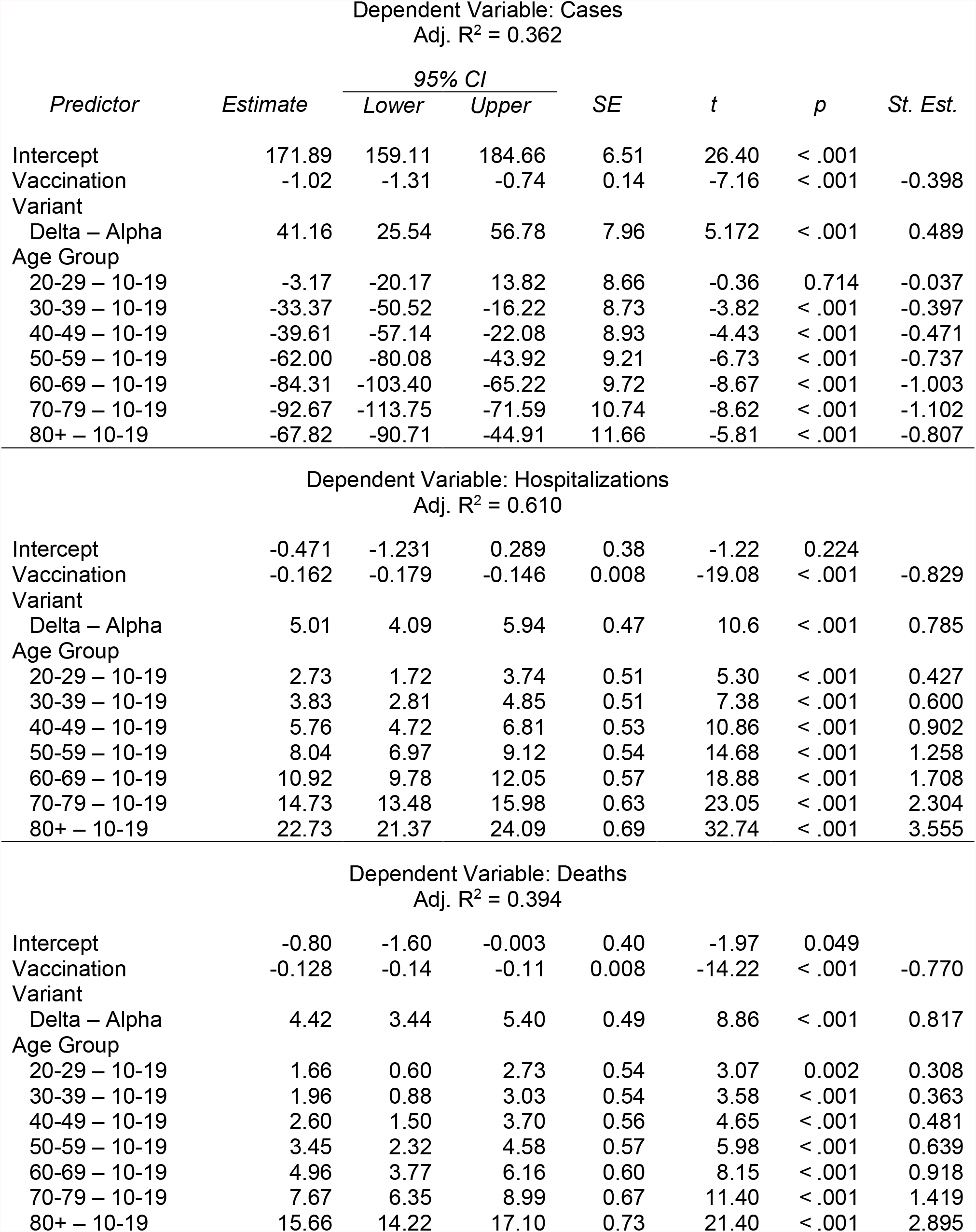
Multiple Linear Regression Results

Dependent Variable: Cases Adj. R<sup>2</sup> = 0.362
| *Predictor* | *Estimate* | *95% CI* |  | *SE* | *t* | *p* | *St. Est.* |
| --- | --- | --- | --- | --- | --- | --- | --- |
|  |  | *Lower* | *Upper* |  |  |  |  |
| Intercept | 171.89 | 159.11 | 184.66 | 6.51 | 26.40 | < .001 |  |
| Vaccination | -1.02 | -1.31 | -0.74 | 0.14 | -7.16 | < .001 | -0.398 |
| Variant |  |  |  |  |  |  |  |
| Delta – Alpha | 41.16 | 25.54 | 56.78 | 7.96 | 5.172 | < .001 | 0.489 |
| Age Group |  |  |  |  |  |  |  |
| 20-29 – 10-19 | -3.17 | -20.17 | 13.82 | 8.66 | -0.36 | 0.714 | -0.037 |
| 30-39 – 10-19 | -33.37 | -50.52 | -16.22 | 8.73 | -3.82 | < .001 | -0.397 |
| 40-49 – 10-19 | -39.61 | -57.14 | -22.08 | 8.93 | -4.43 | < .001 | -0.471 |
| 50-59 – 10-19 | -62.00 | -80.08 | -43.92 | 9.21 | -6.73 | < .001 | -0.737 |
| 60-69 – 10-19 | -84.31 | -103.40 | -65.22 | 9.72 | -8.67 | < .001 | -1.003 |
| 70-79 – 10-19 | -92.67 | -113.75 | -71.59 | 10.74 | -8.62 | < .001 | -1.102 |
| 80+ – 10-19 | -67.82 | -90.71 | -44.91 | 11.66 | -5.81 | < .001 | -0.807 |
| Intercept | -0.471 | -1.231 | 0.289 | 0.38 | -1.22 | 0.224 |  |
| Vaccination | -0.162 | -0.179 | -0.146 | 0.008 | -19.08 | < .001 | -0.829 |
| Variant |  |  |  |  |  |  |  |
| Delta – Alpha | 5.01 | 4.09 | 5.94 | 0.47 | 10.6 | < .001 | 0.785 |
| Age Group |  |  |  |  |  |  |  |
| 20-29 – 10-19 | 2.73 | 1.72 | 3.74 | 0.51 | 5.30 | < .001 | 0.427 |
| 30-39 – 10-19 | 3.83 | 2.81 | 4.85 | 0.51 | 7.38 | < .001 | 0.600 |
| 40-49 – 10-19 | 5.76 | 4.72 | 6.81 | 0.53 | 10.86 | < .001 | 0.902 |
| 50-59 – 10-19 | 8.04 | 6.97 | 9.12 | 0.54 | 14.68 | < .001 | 1.258 |
| 60-69 – 10-19 | 10.92 | 9.78 | 12.05 | 0.57 | 18.88 | < .001 | 1.708 |
| 70-79 – 10-19 | 14.73 | 13.48 | 15.98 | 0.63 | 23.05 | < .001 | 2.304 |
| 80+ – 10-19 | 22.73 | 21.37 | 24.09 | 0.69 | 32.74 | < .001 | 3.555 |
| Intercept | -0.80 | -1.60 | -0.003 | 0.40 | -1.97 | 0.049 |  |
| Vaccination | -0.128 | -0.14 | -0.11 | 0.008 | -14.22 | < .001 | -0.770 |
| Variant |  |  |  |  |  |  |  |
| Delta – Alpha | 4.42 | 3.44 | 5.40 | 0.49 | 8.86 | < .001 | 0.817 |
| Age Group |  |  |  |  |  |  |  |
| 20-29 – 10-19 | 1.66 | 0.60 | 2.73 | 0.54 | 3.07 | 0.002 | 0.308 |
| 30-39 – 10-19 | 1.96 | 0.88 | 3.03 | 0.54 | 3.58 | < .001 | 0.363 |
| 40-49 – 10-19 | 2.60 | 1.50 | 3.70 | 0.56 | 4.65 | < .001 | 0.481 |
| 50-59 – 10-19 | 3.45 | 2.32 | 4.58 | 0.57 | 5.98 | < .001 | 0.639 |
| 60-69 – 10-19 | 4.96 | 3.77 | 6.16 | 0.60 | 8.15 | < .001 | 0.918 |
| 70-79 – 10-19 | 7.67 | 6.35 | 8.99 | 0.67 | 11.40 | < .001 | 1.419 |
| 80+ – 10-19 | 15.66 | 14.22 | 17.10 | 0.73 | 21.40 | < .001 | 2.895 |

## Discussion

This ecological study has several limitations. First, it utilizes data from a single, highly specific population. Second, it considers only fully vaccinated persons. Partially vaccinated persons were not included in analyses^9^, and booster vaccination was not available in Switzerland in the period under consideration. Third, it includes only two COVID-19 variants (i.e., Alpha and Delta). Since the efficacy of currently available vaccines might be reduced with the current dominant variant B.1.1.529 Omicron (Wilhelm et al., preprint [here](https://www.medrxiv.org/content/10.1101/2021.12.13.21267748v2)), this is an important limitation.

That being said, the current study provides some evidence that vaccination is inversely related to COVID-19 cases, hospitalizations, and deaths at the population level, in this case in Switzerland. Further research should include other/several countries, as well as other variants (in particular, Omicron) to confirm or infirm the current findings. While doing so, future research should carefully consider the role of age, variants, and population size [4-5], and use longer observation periods.

## Data Availability

All data produced are available online at https://www.covid19.admin.ch/en/overview

[Note to the Editor: as stated in the Submission guidelines, I have not included preprints in the Reference List. Instead I have cited 4 preprints in the main text and footnotes.]

## Statements and Declarations

The author declares that no funds, grants, or other support were received during the preparation of this manuscript. The author has no relevant financial or non-financial interests to disclose. The author is the only contributor of this manuscript. Ethics approval: not applicable. Consent to participate: not applicable. Consent to publish: not applicable.

## Footnotes

1 That said, see also Harris [4] concerning the most populous counties. For data from France, see Bouanane (preprint: https://orcid.org/0000-0003-2786-2682).

2 In Switzerland, persons who received either two doses of the Moderna (Spikevax) vaccine or Pfizer/BioNTech (Comirnaty) vaccine, or one dose of the Johnson & Johnson (Janssen) vaccine, are deemed fully vaccinated. Switzerland does not use other vaccines.

3 The reason to choose weekly data was that daily data are not available for all our variables of interest by age group and variant.

4 From the end of October 2021, the share of fully vaccinated persons increases only very slowly in Switzerland. To illustrate, from 1 November 2021 to 31 December 2021 (2 months), the share of fully vaccinated persons increased of 3.13 points of percentage, which is low compared to preceding periods (September and October 2021: +9.47; July and August 2021: +16.34; May and June 2021: +25.99 ; March and April 2021: +8.23).

5 Note that the share of Omicron cases was 0% until 18 November 2021. Likewise, the share of any other variant during the period under consideration was never higher than 4.3% (Gamma (P.1) on 24 June 2021). Furthermore, the share of fully vaccinated persons with a booster was 0.01% until 31 October 2021.

6 Switzerland includes data from the Principality of Liechtenstein.

7 The age group from 0 to 9 years old was not included since no more than 0.02% of that population was fully vaccinated during the period under consideration.

8 All tests are two-tailed.

9 The percentage of partially vaccinated persons in Switzerland on the first day and last day of the time period under consideration was 1.08 and 2.30, respectively, with a peak at 15.33 on 15 June 2021.

## Notes

### Competing Interest Statement

The authors have declared no competing interest.

### Funding Statement

This study did not receive any funding.

