## Supplementary Information for "COVID-19 Vaccination is Associated with Decreasing Cases, Hospitalizations, and Deaths Across Age Groups and Variants over 9 months in Switzerland"

**Relationships between Vaccination and Hospitalizations by Age Group and Dominant Variant**

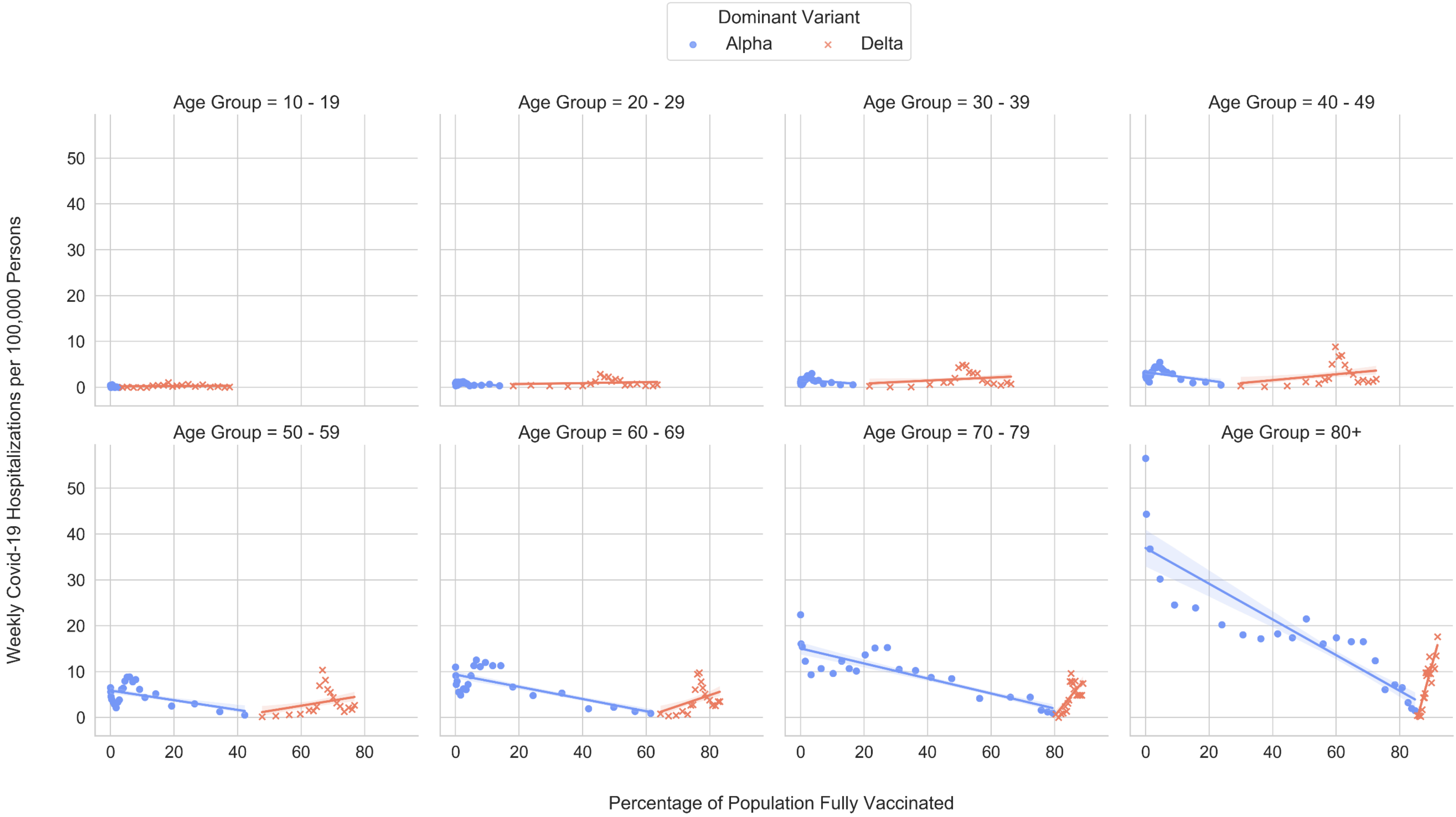

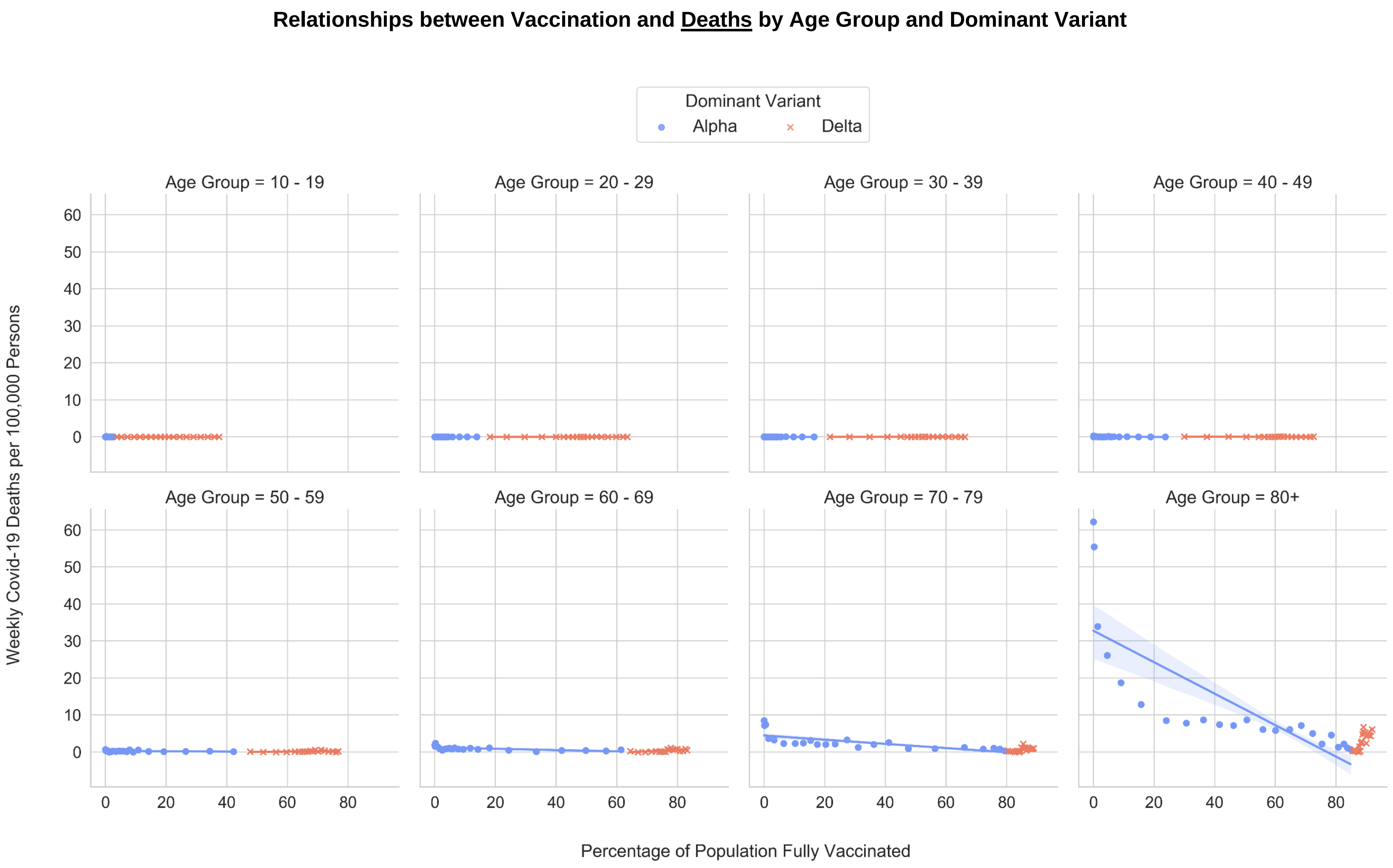
Relationships between Vaccination and Deaths by Age Group and Dominant Variant
Dominant Variant
Alpha
Delta
Age Group = 10 - 19
Age Group = 20 - 29
Age Group = 30 - 39
Age Group = 40 - 49
Age Group = 50 - 59
Age Group = 60 - 69
Age Group = 70 - 79
Age Group = 80+
Weekly Covid-19 Deaths per 100,000 Persons
Percentage of Population Fully Vaccinated
0
10
20
30
40
50
60
0
20
40
60
80
